# Public Perceptions and Engagement in mHealth: A European Survey on Attitudes towards Health Apps Use and Data Sharing

**DOI:** 10.1101/2024.10.10.24315231

**Authors:** Francesco Andrea Causio, Flavia Beccia, Diego Maria Tona, Alessandra Verduchi, Antonio Cristiano, Giovanna Elisa Calabrò, Roberta Pastorino, Carla van El, Stefania Boccia

## Abstract

This study explores public perceptions and engagement in mobile health (mHealth) across eight European countries: Italy, the Netherlands, France, Germany, Spain, Poland, Romania, and Hungary. The focus is on the public’s attitudes toward health app usage and data sharing examined through a cross-sectional survey involving 6,581 participants. The survey revealed that 21.87% of respondents currently use health apps, with 42.71% expressing interest in future use. Regarding data sharing, 52.82% are willing to share health data with healthcare providers, and 25.48% would share data with public and private research institutions. However, concerns about data privacy and security are prevalent, with 63.68% fearing hacking of their data and 72.34% afraid that their data might be used for inappropriate purposes. However, prevalent concerns about data privacy and security emerged, with 72.34% expressing worry about data misuse and 63.68% fearing data hacking.

The study highlights significant generational and geographical differences in mHealth engagement, with older generations displaying a lower adoption level of health apps. Education level emerged as a crucial factor influencing attitudes toward mHealth, with those having tertiary education more likely to use health apps and demand transparency. These findings underscore the need for targeted strategies to enhance digital literacy, ensure data privacy, and promote equitable access to mHealth technologies across Europe.

## Introduction

Mobile Health (mHealth) is a transformative approach leveraging mobile devices, such as smartphones and wearables, to enhance medical and public health practices. Patient education apps, medication reminders, and telemedicine apps have seen a significant rise in recent years, supporting disease management and promoting wellness^1^.

Similarly, Electronic Health Records (EHRs) are increasingly adopted in healthcare^2^. These digital records of patients’ health information are intended to be shared across different healthcare settings, offering a broader view of a patient’s care^34^. Integrated with EHRs are patient portals: these are secure online applications that allow patients access to their health information on medical records, schedule appointments, and securely communicate with their health professionals^5^.

Both mHealth and EHRs depend on patients’ consent for the sharing of health data, enabling physicians and third parties to access their health information. User-centric health data sharing safeguards users’ privacy while also fostering trust^6^. According to Kim et al.^7^, users support health data sharing, provided that there is transparency in data control, access, and purpose of use. Buhr et al. found a high potential for adopting research-oriented apps among smartphone users, especially when data is handled by state-funded or governmental institutions^8^.

The primary use of health data pertains to its sharing for individual benefit, distinguishing it from the secondary use of aggregated health data by various entities such as researchers and policymakers^9^. Health data, including genomic data, plays a crucial role in enabling researchers to uncover new preventive interventions, therapies and understand determinants of health and disease. Unsurprisingly, collective involvement and shared participation of individuals leads to improved research outcomes^10^.

Previous studies have addressed some of the determinants and predictors involved in health app adoption and data sharing, in individual countries or small samples^11^. The “European network staff eXchange for integrating precision health in the Health Care Systems” (ExACT) project disseminated a cross-sectional survey involving 6,581 participants from eight European countries: Italy, the Netherlands, France, Germany, Spain, Poland, Romania, and Hungary, investigating their attitudes towards personalized medicine, health apps, and health data sharing. This research builds on earlier studies, including the Your DNA Your Say (YDYS) project and an Italian survey addressed to the general public^12,13^. However, these studies focused on health data sharing, leaving mHealth and EHRs out of their scope. This study explores the European population’s comprehension and perspectives regarding health apps and health data sharing, with a particular focus on the influence of different social generations on the adoption of health apps^14, 15^.

## Methods

### Questionnaire

A 37-question web questionnaire (available in the Supplementary file) was designed and divided into four primary sections: Module A, Knowledge and attitudes about Personalized Medicine; Module B, Genomic and health data sharing and use; Module C, Governance; Module D, Needs of users. Researchers hired the private company YouGov to disseminate the survey using YouGov’s polling platform (https://yougov.co.uk/about/panel-methodology). The survey was distributed over a period of two weeks in April 2023. Participants for the YouGov survey were selected based on demographic information to reflect the population distribution of their respective countries in terms of gender, age, and education level. Ethical approval for this study was obtained from the Policlinico Universitario ‘Agostino Gemelli’ Ethics Committee, Rome (ID 5047) and Amsterdam UMC (reference 2022.0214). The survey’s design and delivery methods have been comprehensively described in a previously published manuscript^16^. For the specific purposes of this study, we restricted the analysis on the six questions specifically dedicated to health apps (Question 16 to 21).

The following methods section is dedicated to the data preprocessing and analysis relevant to the specific objective of the study.

### Development of composite indicators

Following the methodology previously used in other analogous surveys^13,17,18^, we built four specific indicators: “Current Health App Usage”, “Indicator for positive attitudes towards using health apps and data sharing”, “Higher Information Requests” and “Distrustful Attitude towards Health Apps”. These indicators were built to measure respondents’ current usage of health apps, their composite attitudes towards the use of health apps and data sharing, transparency measures required for health apps use and perceived fears in health apps use.

The “Current Health App Usage” indicator was established based on question 16, categorizing respondents who answered “Yes, I already use such apps” as “Current user” and those who provided any other response as “Non-user” (Table 1).

**Table 1.** Components of the indicators. In the Questions column, the italic number in brackets indicates the score assigned to each question.

| Indicator | Questions | Coding |
| --- | --- | --- |
| Current Health App Usage | Would you be interested in using personal health care apps monitoring your health (e.g. heart rate, exercise, response to medications)? | Yes, I already use such apps → Current User<br>Yes, I might use such apps in the future / No, I am not interested / Don't know → Non-user |
| Indicator for positive attitudes towards using health apps and data sharing | Would you be interested in using personal health care apps monitoring your health (e.g. heart rate, exercise, response to medications)? (0-1)<br>If it were possible in your country to share the data from your personal health apps with your medical record/health portal so your health care provider or doctor would have access, would you be willing to share your data? (0-1)<br>And would you be willing to share the data from your personal health apps with research institutes or biobanks? (0-1) | The cut-off for positive or negative attitudes towards using health apps and data sharing was set at 67%<br>- Positive attitudes if $> 2/3$<br>- Negative attitudes if $\leq 2/3$ |
| Higher Information Requests | What information would you like to be provided to you before using these health apps? Please select all that apply. (0-7) | 0-2 requested information → Low requested information<br>3-7 requested information → High requested information |
| Distrustful Attitude towards Health Apps | Which, if any, of the following do you perceive as potential risks related to sharing your data in health apps? Please select all that apply. (0-8) | 0-2 perceived potential risks → Low potential risks perceived<br>3-8 perceived potential risks → High potential risks perceived |

The “Indicator for positive attitudes towards using health apps and data sharing” indicator was built upon multiple questions: question 16 (scoring 0-1 points based on participants’ current use of health apps assigning 1 point to those responded “Yes, I might use such apps in the future” and 0 points to all other responses), question 18 (scoring 0-1 points depending on participants’ expressed interest in sharing their health apps data with their patient portal for their healthcare provider’s access assigning 1 point to those responded “Yes” and 0 points to responses “No” or “Don’t know”), and question 19 (scoring 0-1 points depending on participants’ interest in sharing their health data with biobanks and research institutions assigning 1 point to those who responded “Yes”, regardless of the institutions type, or “Both”, and 0 points to responses “No” or “Don’t know”). The maximum achievable value for this indicator is 3. Respondents with an indicator value of ≥2/3 (67%) were considered to have a positive attitude towards health app use and data sharing, while those with a lower value were considered to have a negative attitude (Table 1).

An indicator related to the amount of information requested prior to using health apps named “Higher Information Requests”, was built using Question 17 “What information would you like to be provided before using these health apps? Please select all that apply”. Respondents could choose up to eight different options, including one open option, or state that they would not request information to use health apps. Because the ‘Other’ option had significantly fewer respondents compared to other options, it was not included in this analysis (Table 1). The “Higher Information Requests” indicator was created as a binary variable, as follows: using a 25% cut-off, respondents requesting 0, 1, or 2 types of information were categorized as requesting low information whereas respondents requesting 3 to 7 types of information were categorized as requesting high information.

An indicator called “Distrustful Attitude towards Health Apps”, was built using Question 20, “Which, if any, of the following do you perceive as potential risks related to sharing your data in health apps?”. Respondents could choose up to eight different options, including one open option, or state that they did not perceive any risks from using health apps. Due to fewer respondents selecting the ‘Other’ option compared to the other choices, it was excluded from this analysis (Table 1). The “Distrustful Attitude towards Health Apps” indicator was created as a binary variable, and built as follows: using a 25% cut-off, respondents requesting 0, 1, or 2 types of information were categorized as perceiving low risks; conversely, respondents perceiving 3 to 8 risks were categorized as perceiving high risks.

### Data Analysis

The statistical analysis was performed using STATA 18.0 software (Stata Corporation, College Station, TX, USA).

We applied descriptive statistics to analyze the results, using absolute frequencies and percentages for categorical variables and means and standard deviations (SD) for continuous variables.

To investigate the association between the indicators detailed in the previous section and gender, social generation, geographical region, and education, we applied multivariable logistic regression models^19^. Social generation categories were determined based on the following cutoffs: 1928-1945 for the Silent Generation, 1946-1964 for Baby Boomers, 1965-1980 for Generation X, 1981-1996 for Millennials, and 1997-2005 for Generation Z, in line with the cut offs of the Pew Research Center^20^. Education levels were dichotomized as having achieved tertiary education or not. The geographical area was categorized as follows: Eastern Europe (Poland, Hungary, and Romania); Southern Europe (Italy and Spain), and Central Europe (the Netherlands, Germany, and France). Each variable was examined by univariable analysis and was included in the multivariable logistic model when the P value was < 0.15. The influence of the independent variables on each binary outcome investigated was expressed as odds ratios (ORs) and 95% confidence interval (CI).

## Results

### Demographics

The total number of respondents was 6,581, with females comprising 52.55% of the sample (n= 3,458). The age distribution ranged from 18 to 89 years (mean = 48.5 years, median = 49 years, standard deviation = 15.96 years). The majority of participants belonged to Generation X, constituting 31.15% (n = 2,050) of the total. Baby Boomers followed closely, accounting for 29.69% (n = 1,954), while Millennials comprised 26.55% (n = 1,747) of the sample. Generation Z, the youngest generation in the sample, represented 10.86% (n = 715) of participants, whereas the Silent Generation, the oldest group, constituted a smaller proportion of 1.75% (n = 115). Participants hailed from eight distinct European nations. Of these, Central Europe — including France, Germany, and the Netherlands — represented the largest segment, with 46.03% (n=3,029) of the total respondents. Eastern Europe, including Hungary, Poland, and Romania, comprised 23.20% (n=1,527) of the cohort, while Southern Europe, with Italy and Spain, comprised 30.77% (n=2,025). Specifically, 1,022 respondents were from Italy, 1,012 from the Netherlands, 1,009 from Germany, 1,003 from Spain, 1,000 from France, 510 from Hungary, 509 from Poland, and 508 from Romania. Educational levels among the respondents were diverse, grouped according to whether they had achieved tertiary education (37.91%, n=2,495) or not (62.09%, n=4,086). The participants’ demographics are detailed in Table 2.

**Table 2.** Demographic and general information.

| Demographic information (N=6,581). |  | No. | % |
| --- | --- | --- | --- |
| Gender |  |  |  |
|  | Male | 3,123 | 47.5 |
|  | Female | 3,458 | 52.5 |
| Age (years) |  | Average = 48.5 | SD = 16.0 |
| Social Generation |  |  |  |
|  | Generation Z | 715 | 10.86 |
|  | Millennials | 1,747 | 26.55 |
|  | Generation X | 2,050 | 31.15 |
|  | Baby Boomers | 1,954 | 29.69 |
|  | Silent Generation | 115 | 1.75 |
| Achieved tertiary education |  |  |  |
|  | No | 4,086 | 62.1 |
|  | Yes | 2,495 | 37.9 |
| Country |  |  |  |
|  | France | 1,008 | 15.3 |
|  | Germany | 1,009 | 15.4 |
|  | Hungary | 510 | 7.7 |
|  | Italy | 1,022 | 15.5 |
|  | Netherlands | 1,012 | 15.4 |
|  | Poland | 509 | 7.7 |
|  | Romania | 508 | 7.7 |
|  | Spain | 1,003 | 15.3 |
| Geographic area |  |  |  |
|  | Central Europe | 3,029 | 46.03 |
|  | Eastern Europe | 1,527 | 23.20 |
|  | Southern Europe | 2,025 | 30.77 |

### Interest in using health apps

When assessing interest in personal health care applications for health monitoring, it was shown that 21.87% of the respondents were already using such apps, whereas 42.71% expressed interest in future use despite not currently engaging with these technologies. Conversely, 23.84% indicated no interest in health monitoring apps, and 11.58% were uncertain (Table 3).

**Table 3.** Interest in using health apps for health monitoring and data sharing.

| Would you be interested in using personal health care apps monitoring your health (e.g., heart rate, exercise, response to medication)? | No. | % |
| --- | --- | --- |
| Yes, I already use such apps | 1,439 | 21.87% |
| Yes, I might use such apps in the future | 2,811 | 42.71% |
| No, I am not interested | 1,569 | 23.84% |
| Don't Know | 762 | 11.58% |
| If it were possible in your country to share the data from your personal health apps with your medical record/health portal so your health care provider or doctor would have access, would you be willing to share your data? | No. | % |
| Yes | 3,476 | 52.82% |
| No | 1,514 | 23.01% |
| Don't know | 1,591 | 24.18% |
| Would you be willing to share the data from your personal health apps <u>with research institutes or biobanks</u>? | No. | % |
| Yes, only public | 1,469 | 22.32% |
| Yes, only private | 591 | 8.98% |
| Yes, Both | 1,677 | 25.48% |
| No | 1,409 | 21.41% |
| Don't know | 1,435 | 21.81% |

Among the different countries considered, Spain stands out with the highest percentage of current users of health apps at 29.51%, whereas France had the lowest (14.29%). Italy featured with the highest percentage of respondents who might be interested in using health apps in the future, at 51.17%, whereas the Netherlands had the lowest (32.04%). Those not interested in using health apps were the most represented category among respondents from Germany (36.47%), with Hungary having the least uninterested respondents (14.51%) (Supplementary Material).

### Sharing Data from Health Apps with Patient Portals and Research Institutes or Biobanks

When asked whether they would share the data from their personal health apps with their medical record or health portal for their healthcare provider or physician to have access, most (52.82%) responded affirmatively; in contrast, 23.01% of the respondents would not be willing to share their data, and almost one in four (24.18%) remained uncertain (Table 3).

Romania (62.40%) had the most respondents open to sharing their data, and France (29.07%) showed the most unwilling respondents (Supplementary Material).

Respondents’ willingness to share personal health app data with research institutes or biobanks varies depending on whether it is a public or private institution. 25.48% are willing to share with both public and private organizations; conversely, 22.32% would share their data only with public research institutions, and 8.98% would share exclusively with private entities (Table 3). Different countries show fluctuating levels of willingness with Hungary having the highest proportion of respondents willing to share with both public and private institutions (38.04%) but the least willing to share with public institutions only (16.27%). When it comes to sharing data with private institutions only, Germany stands as the country with the least willing to share (6.74%), whereas Poland has the highest values (11.59%) (Supplementary Material).

### Requested information and Transparency for the Use of Health Apps

When investigating information deemed relevant before using health apps, the most emphasized detail was data usage and access (52.59%) and a clear description of the app’s purposes (46.06%) followed by the right to delete personal data (44.02%) and by details on privacy policies (43.69%) and data storage (43.29%). Information on data portability to other devices or platforms and the release on information about the developers were less frequently considered relevant (29.11% and 23.10%, respectively) (Table 4).

**Table 4.** Requested information related to the use of health apps (multiple-select questions)

| Requested Transparency Disclosures | No. | % |
| --- | --- | --- |
| Data Usage and Access | 3,461 | 52.59% |
| Description of the App's Purposes | 3,031 | 46.06% |
| Right to Delete Personal Data | 2,897 | 44.02% |
| Privacy Policies Details | 2,875 | 43.69% |
| Data Storage Information | 2,849 | 43.29% |
| Data Portability to Other Devices/Platforms | 1,916 | 29.11% |
| Information about the Developers | 1,520 | 23.10% |
| Other Information | 93 | 1.41% |

### Perceived potential risks Connected to Health Apps

The most perceived risk related to sharing data in health apps is data misuse (72.34%), including data being used for personal identification, for discriminatory purposes by the government, for health-related stigma (e.g., on the workplace or in obtaining an insurance) or for commercial gain. Following closely is the perception of data being hacked (63.68%) and being used for unauthorized purposes or leading to identity theft. Finally, perceived risks included data being wrongfully reported or, in the case of genetic data, being linked to a crime (39.61%) (Table 5).

**Table 5.** Perceived potential risks in health app use (multiple-select questions)

| Number of citizens perceiving data hacking, being wrongfully reported, or being misused, in connection to the use of health apps use |  |  |
| --- | --- | --- |
| Category | No. | % |
| Data Misuse | 4,761 | 72.34% |
| Data Hacking | 4,191 | 63.68% |
| Data being Wrongfully Reported | 2,607 | 39.61% |

### Logistic regression

#### Predictors of Current Health App Usage

The probability of using health apps is negatively associated with older social generations. Specifically, individuals from older generations are less likely to use health apps compared to Generation Z. This trend becomes more pronounced with each successive older generation: Generation X (OR 0.74, 95% CI 0.60-0.90, p = 0.003), Baby Boomers (OR 0.54, 95% CI 0.44-0.66, p < 0.001), and the Silent Generation (OR 0.47, 95% CI 0.27-0.81, p = 0.007). Geographical differences also play a significant role. Compared to respondents from Central Europe, those from Eastern Europe are more likely to use health apps (OR 1.47, 95% CI 1.26-1.70, p < 0.001), as are respondents from Southern Europe (OR 1.40, 95% CI 1.22-1.61, p < 0.001). Additionally, individuals with tertiary education are more likely to use health apps (OR 1.41, 95% CI 1.25-1.59, p < 0.001) (Table 6).

**Table 6.** Predictors of Current Health App Usage, Positive Attitudes towards Health Apps Use and Data Sharing, Higher Information Requests, and a Distrustful Attitude towards Health Apps in Europe.

| **Predictors of Current Health App Usage** |  |  |
| --- | --- | --- |
| Predictor | OR (95% CI) | P-value |
| Gender (*reference = Male*) |  |  |
| Female | 1.12 (0.99 – 1.26) | 0.068 |
| Social Generation (*reference = Generation Z*) |  |  |
| Millennials | 0.87 (0.71 – 1.06) | 0.158 |
| Generation X | 0.74 (0.60 – 0.90) | **0.003** |
| Baby Boomers | 0.54 (0.44 – 0.66) | **< 0.001** |
| Silent Generation | 0.47 (0.27 – 0.81) | **0.007** |
| Geographical area (*reference = Central Europe*) |  |  |
| Eastern Europe | 1.47 (1.26 – 1.70) | **< 0.001** |
| Southern Europe | 1.40 (1.22 – 1.61) | **< 0.001** |
| Education Level (*reference = Education level below tertiary*) |  |  |
| Tertiary education | 1.41 (1.24 – 1.58) | **< 0.001** |
| **Predictors of Positive attitudes towards using Health Apps and Data Sharing** |  |  |
| Predictor | OR (95% CI) | P-value |
| Gender (*reference = Male*) |  |  |
| Female | 0.82 (0.73 – 0.92) | **< 0.001** |
| Social Generation (*reference = Generation Z*) |  |  |
| Millennials | 0.95 (0.78 – 1.17) | 0.690 |
| Generation X | 0.90 (0.74 – 1.09) | 0.275 |
| Baby Boomers | 1.13 (0.92 – 1.38) | 0.22 |
| Silent Generation | 2.38 (1.36 – 4.16) | **0.002** |
| Geographical area (*reference = Central Europe*) |  |  |
| Eastern Europe | 1.72 (1.48 – 1.98) | **< 0.001** |
| Southern Europe | 1.99 (1.73 – 2.27) | **< 0.001** |
| Education Level (*reference = Education level below tertiary*) |  |  |
| Tertiary education | 1.21 (1.07 – 1.36) | **< 0.001** |
| **Predictors of Higher Information Requests** |  |  |
| Predictor | OR (95% CI) | P-value |
| Gender (*reference = Male*) |  |  |
| Female | 1.15 (1.04-1.27) | **< 0.004** |
| Social Generation (*reference = Generation Z*) |  |  |
| Millennials | 1.12 (0.94 – 1.35) | 0.189 |
| Generation X | 1.57 (1.31 – 1.87) | < **0.001** |
| Baby Boomers | 2.11 (1.77 – 2.52) | < **0.001** |
| Silent Generation | 1.61 (1.07 – 2.40) | **0.020** |
| Geographical area (*reference = Central Europe*) |  |  |
| Eastern Europe | 1.45 (1.28 - 1.65) | < **0.001** |
| Southern Europe | 1.18 (1.05 – 1.33) | **0.004** |
| Education Level (*reference = Education level below tertiary*) |  |  |
| Tertiary education | 1.61 (1.45 – 1.78) | < **0.001** |
| **Predictors of a Distrustful Attitude towards Health Apps** |  |  |
| Predictor | OR (95% CI) | P-value |
| Gender (*reference = Male*) |  |  |
| Female | 1.13 (1.02-1.24) | **0.017** |
| Social Generation (*reference = Generation Z*) |  |  |
| Millennials | 1.31 (1.09 – 1.56) | <**0.001** |
| Generation X | 1.89 (1.59 – 2.26) | < **0.001** |
| Baby Boomers | 2.45 (2.05 – 2.92) | < **0.001** |
| Silent Generation | 2.19 (1.47 – 3.27) | < **0.001** |
| Education Level (*reference = Education level below tertiary*) |  |  |
| Tertiary education | 1.49 (1.34 – 1.65) | < **0.001** |

#### Predictors of Positive Attitudes towards using Health Apps and Data Sharing

Females show lower odds of displaying positive attitudes than males (OR 0.82, 95% CI 0.73-0.92, p < 0.001). The Silent Generation is more likely to have positive attitudes towards the use of health apps and data sharing (OR 2.38, 95% CI 1.36 – 4.16, p = 0.002). Respondents from Eastern Europe have significantly higher odds of displaying positive attitudes towards health app use and data sharing than those from Central Europe (OR 1.72, 95% CI 1.48-1.98, p < 0.001). Similarly, respondents from Southern Europe have higher odds than the Central European reference, with an OR of 1.99 (95% CI 1.73-2.27, p < 0.001). Having tertiary education is associated with higher odds of displaying positive attitudes, with an OR of 1.21 (95% CI 1.07-1.36, p < 0.001). The results for this section are reported in Table 6.

#### Predictors of Higher Information Requests

Compared to Generation Z, individuals from older generations were more likely to request information, with the likelihood increasing progressively from Generation X (OR 1.57, 95% CI 1.31-1.87, p < 0.001) to Baby Boomers (OR 2.11, 95% CI 1.77-2.52, p < 0.001), and then slightly decreasing in the Silent Generation (OR 1.61, 95% CI 1.07-2.40, p < 0.001). Females are more likely to request additional information compared to males (OR 1.15, 95% CI 1.04-1.27, p = 0.004). Furthermore, individuals with higher education levels are more likely to request more information before using health apps (OR 1.61, 95% CI 1.45-1.78, p < 0.001) (Table 6).

#### Predictors of a Distrustful Attitude towards Health Apps

Compared to Generation Z, individuals from older generations were more likely to have a distrustful attitude towards health apps. Specifically, Baby Boomers had an OR of 2.45 (95% CI 2.05-2.92, p < 0.001) and the Silent Generation had an OR of 2.19 (95% CI 1.47-3.27, p < 0.001). Females had an OR of 1.13 (95% CI 1.02-1.24, p=0.017), and individuals having achieved tertiary education had an OR of 1.49 (95% CI 1.34-1.65, p < 0.001).

## Discussion

Our study provides a comprehensive overview of public perceptions and engagement in mobile health (mHealth) technologies across eight European countries. Through a cross-sectional survey involving 6,581 participants, we investigated attitudes towards health app usage and data sharing, as well as their perceived potential risks and transparency disclosures.

### Generational Differences in Technology Adoption

Different social generations exhibit varying levels of engagement with new technologies. Younger generations demonstrate greater enthusiasm for adopting health apps, reflecting a broader trend in digital technology adoption. Wellness apps have garnered significant traction among younger demographics, with research indicating that younger, healthier individuals are more inclined to share certain types of health information, such as weight, diet, and lifestyle data^21^. However, concerns about sharing disease-specific and genetic information remain pronounced.

In contrast, health app usage among older generations, including Baby Boomers, remains considerably lower, highlighting a significant underutilization of digital health potential, especially in managing chronic conditions, which are more prevalent among older demographics^22^.

However, our findings suggest that members of the Silent Generation, typically older adults, exhibit a greater propensity to embrace health apps compared to Generation Z. This could be attributed to an increasing awareness of health concerns among older adults, leading to a greater perceived benefit in utilizing health apps and sharing health data to maintain well-being. Additionally, older generations also demonstrate an increased perception of potential risks associated with health app usage, which underscores the importance of addressing these concerns through enhanced education and reassurance regarding data privacy and security^23^.

### Educational and Geographical disparities

Citizens with a tertiary level of education use health apps more frequently than those without, confirming the association between education and health app usage previously found in literature^24^. Individuals with higher educational levels are also more likely to share their health app data with health portals. These trends could be explained by greater knowledge of the benefits of using these applications and better digital literacy^25^.

Achieving tertiary education is a positive predictor of using health apps and is also strongly associated with a generally positive attitude towards them and data sharing. This suggests that the general public’s positive attitudes towards health apps and data sharing are positively influenced by their confidence and capability in handling the information they are given. Informing citizens about the possibilities connected to health apps and data sharing, along with thorough description of any attached strings, will play a key role in fostering positive attitudes.

We previously observed that Eastern and Southern Europe have consistently higher knowledge levels and positive attitudes towards data sharing when compared to Central Europe^26^. This heterogeneity in Europe persists in the adoption of health apps, with respondents in Eastern and Southern Europe being more likely to use health apps than those in Central Europe.

### Data Privacy and Security Concerns

The survey results showed that the information citizens seek before using health apps includes disclosures on how the data will be used, who has access to it, and the general purposes of the app. Concurrently, data being hacked or used for commercial purposes were the most common perceived risks.

Specifically, respondents appear to be more reluctant if the recipient is a for-profit researcher, company, or government; existing literature suggests this is related to a lack of information on how the data will be used and how privacy will be guaranteed^27^. A public consultation from the European Commission in 2014 led to the realization of the “Privacy code of conduct on mobile health apps” to promote trust among users^28^. Despite these efforts, some health apps on the market still have poor data privacy, sharing, and security standards^29^.

Most respondents were interested in sharing data from health apps with patient portals and research institutes or biobanks. Although this willingness can be seen as a positive development in the integration of digital health technologies into mainstream healthcare, it’s worth noticing that a substantial portion of respondents were unwilling to share their data. These findings highlight concerns or reservations among individuals regarding the privacy and security of their health data^30^. The data also suggest that individuals are discerning about sharing health data with different types of institutions. Male participants showed a greater willingness to share, and those with tertiary education were less prone to share exclusively with private entities. Country-wide variations were pronounced, with Hungary standing out as the country with the highest openness to sharing with both public and private institutions.

### Future perspectives

The digital transformation of healthcare systems, accelerated by the COVID-19 pandemic, underscores the significance of health data as a pivotal asset and places digital health solutions at the forefront of modern healthcare^31^. mHealth solutions, facilitating ongoing communication between patients and their healthcare providers, empower physicians to deliver care not only in traditional face-to-face settings but also remotely^32^.

However, alongside the opportunities presented by mHealth, there are inherent risks, such as digital exclusion, misinformation proliferation, and the promotion of unhealthy behaviors^33^. Systematic assessment and evaluation of the impact of digital health services is needed, as evidence on their effectiveness and cost-effectiveness remains limited. Evaluations should consider the broad goals of health systems, including quality, efficiency, equity, and patient empowerment. A European repository of evaluation frameworks, methods, and evidence could facilitate knowledge exchange and continuous improvement. Given the rapid pace of innovation, flexible and adaptable evaluation approaches are required, along with efforts to align decentralized decision-making with overall health system objective^34^.

Patient engagement is an important catalyst for the effectiveness of health apps and is associated with better health outcomes, especially in managing chronic conditions such as diabetes and hypertension^35,36^. Previous literature shows that feedback on one’s progress and the ability to set goals and receive rewards are regarded as very important^37^. The significant association of increasing age and education levels with increased requests for information and higher perceived potential risks underlines the need for transparency when disclosing information on health app functioning, including when Artificial Intelligence algorithms are implemented.

## Study Limitations

This study should be considered in light of some limitations.

This survey was administered on an online platform; hence some respondents, which might have introduced a level of negligence in respondents’ completion of the survey. Additionally, participation in the survey was voluntary, potentially resulting in self-selection bias. Consequently, the perspectives of those who chose to participate may differ from those who opted not to participate, leading to a skewed representation of opinions.

Furthermore, while efforts were made to weight the selected sample according to the distribution of gender, age, and education level in the identified countries, variations in representativeness across different categories may have compromised the validity of comparisons.

## Conclusion

The study provides a comprehensive overview of the general public’s attitudes towards health app usage and data sharing in Europe, highlighting diverse perceptions and engagement levels across Central, Southern, and Eastern Europe. It underscores the growing role of health apps in healthcare and the crucial influence of demographic factors like age, education, and geographic location on their adoption and use. The research reveals a notable willingness among the public to engage with mHealth technologies, especially among those with higher educational levels and older generations alike. However, it also underscores significant concerns regarding data privacy and security, which are top priorities for users. These findings call for a multifaceted approach to address these concerns, enhance digital health literacy, and ensure equitable access to mHealth. Policymakers and healthcare providers must prioritize the development of robust, transparent, and user-friendly digital health platforms that respect user privacy and foster trust. By addressing these key areas, the potential of mHealth to revolutionize healthcare delivery and patient outcomes can be fully realized, paving the way for more personalized, efficient, and accessible healthcare systems across Europe.

## Data Availability

All data produced in the present study are available upon reasonable request to the authors

## Acknowledgments

We thank the respondents to the survey who took the time to answer the questions on personalized medicine. We thank the Italian Ministry of Health for supporting the activities of the National Center for Disease Prevention and Control (CCM) projects related to genomics and omics sciences, including implementing our survey.

## Declarations

### Ethics approval and consent to participate

Ethical approval for this study was obtained from the Policlinico Universitario ‘Agostino Gemelli’ Ethics Committee in Rome (ID 5047) and Amsterdam UMC (reference 2022.0214). All participants were informed about the purpose of the study and their rights as participants, including the voluntary nature of their involvement. Consent was obtained prior to participation, ensuring that respondents understood their data would be used for research purposes while maintaining confidentiality and anonymity. Participants were also made aware of their right to withdraw from the study at any time without any consequences.

