## Supplementary Material for "Public Perceptions and Engagement in mHealth: A European Survey on Attitudes towards Health Apps Use and Data Sharing"

***Table 1. Public interest in using health apps by country. Percentage values indicate the relative frequency within the country.***

| Would you be interested in using personal health care apps monitoring your health (e.g., heart rate, exercise, response to medication)? | | | | | | | | | |
|---|---|---|---|---|---|---|---|---|---|
| | FR | DE | HU | IT | NL | PL | RO | ES | EU |
| Uses health apps | 144 **14.29 %** | 162 16.06 % | 144 28.24 % | 195 19.08 % | 255 25.20 % | 102 20.04% | 141 27.76% | 296 **29.51%** | 1439 21.87% |
| Interested but does not use health apps | 449 44.54 % | 357 35.38 % | 225 44.12 % | 523 **51.17 %** | 324 **32.04 %** | 251 49.31% | 245 48.23% | 437 43.57% | 2811 42.71% |
| Not interested | 291 28.87 % | 368 **36.47 %** | 74 **14.51 %** | 172 16.83 % | 325 32.11 % | 91 17.88% | 76 14.96% | 172 17.15% | 1569 23.84% |
| Doesn't Know | 124 12.30 % | 122 12.09 % | 67 **13.14 %** | 132 12.92 % | 108 10.67 % | 65 12.77% | 46 **9.06%** | 98 9.77% | 762 11.58% |
| Total | 1008 100% | 1009 100% | 510 100% | 1022 100% | 1012 100% | 509 100% | 508 100% | 1003 100% | 6581 100% |

FR= France DE = Germany HU = Hungary IT = Italy NL = Netherlands PL = Poland RO =Romania ES = Spain TOT = Total. Bold values indicate the highest and lowest values in the range.

***Table 2. Citizens' willingness to share health data with patient portals. Percentage values indicate the relative frequency within the country.***

| If it were possible in your country to share the data from your personal health apps with your medical record/health portal so your health care provider or doctor would have access, would you be willing to share your data? | | | | | | | | | |
|---|---|---|---|---|---|---|---|---|---|
| | FR | DE | HU | IT | NL | PL | RO | SP | EU |
| Yes | 456 45.24 % | 445 **44.10 %** | 296 58.04 % | 614 60.08 % | 562 55.53% | 230 45.19% | 317 **62.40 %** | 556 55.43 % | 3476 52.82% |
| No | 293 29.07 % | 329 **32.61 %** | 90 17.65 % | 178 17.42 % | 232 22.92% | 109 21.41% | 81 **15.94 %** | 202 20.14 % | 1514 23.01% |
| Don't know | 253 25.69 % | 235 23.29% | 124 24.31 % | 230 22.50 % | 218 **21.54 %** | 170 **33.40 %** | 110 21.65% | 245 24.43 % | 1591 24.18% |
| Total | 1008 100% | 1009 100% | 510 100% | 1022 100% | 1012 100% | 509 100% | 508 100% | 1003 100% | 6581 100% |

FR= France DE = Germany HU = Hungary IT = Italy NL = Netherlands PL = Poland RO =Romania ES = Spain TOT = Total. Bold values indicate the highest and lowest values in the range.

***Table 3. Citizen's willingness to share health data with research institutes and biobanks. Percentage values indicate the relative frequency within the country.***

| Would you be willing to share the data from your personal health apps with research institutes or biobanks? | | | | | | | | | |
|---|---|---|---|---|---|---|---|---|---|
| | FR | DE | HU | IT | NL | PL | RO | SP | EU |
| Yes, only public | 207 20.54 % | 194 19.23% | 83 **16.27 %** | 278 **27.20 %** | 217 21.44% | 102 20.04% | 137 26.97% | 251 25.02% | 1,469 22.32 % |
| Yes, only private | 102 10.12 % | 68 **6.74%** | 57 11.18% | 69 6.75% | 110 10.87% | 59 **11.59 %** | 41 8.07% | 85 8.47% | 591 8.98% |
| Yes, Both | 200 19.84 % | 224 22.20% | 194 **38.04 %** | 306 29.94% | 187 **18.48 %** | 118 23.18% | 133 26.18% | 315 31.41% | 1,677 25.48 % |
| No | 249 24.70 % | 314 **31.12 %** | 70 13.73% | 154 15.07% | 104 20.43% | 104 20.43% | 96 18.90% | 134 **13.36 %** | 1,409 21.41 % |
| Don't know | 250 **24.80 %** | 209 20.71% | 106 20.78% | 215 21.04% | 126 24.75% | 126 24.75% | 101 **19.88 %** | 218 21.73% | 1,435 21.81 % |
| Total | 1,008 100% | 1,009 100% | 510 100% | 1,022 100% | 1,012 100% | 509 100% | 508 100% | 1,003 100% | 6,581 100% |

FR= France DE = Germany HU = Hungary IT = Italy NL = Netherlands PL = Poland RO =Romania ES = Spain TOT = Total. Bold values indicate highest and lowest values in the range.

***Table 4. Number of required transparency disclosures.***

| Number of citizens asking for transparency disclosures (N = cumulative number of transparency disclosures asked for). | | |
|---|---|---|
| | Absolute value | Relative value |
| N=0 | 1035 | 15.73% |
| N=1 | 1423 | 21.62% |
| N=2 | 893 | 13.57% |
| N=3 | 947 | 14.39% |
| N=4 | 632 | 9.60% |
| N=5 | 537 | 8.16% |
| N=6 | 424 | 6.44% |
| N=7 | 685 | 10.41% |
| N=8 | 5 | 0.08% |

*Table 5. Perceived potential risks in health app use.*

| Number of citizens perceiving N risks connected to health app use (data hacked, data used for unauthorized purposed, data used for commercial gains, being identified, identity theft, discrimination by the government, discrimination in the workplace, being accused of a crime, unspecified others) | | |
|---|---|---|
| | Absolute value | Relative value |
| N=0 | 1034 | 16.17% |
| N=1 | 1215 | 18.46% |
| N=2 | 984 | 14.95% |
| N=3 | 1087 | 16.52% |
| N=4 | 730 | 11.09% |
| N=5 | 498 | 7.57% |
| N=6 | 367 | 5.58% |
| N=7 | 284 | 4.32% |
| N=8 | 351 | 5.33% |
| N=9 | 1 | 0.02% |
